# BIOIMAGING AND COMPARATIVE GENOMICS UNCOVER PERSISTENCE-ASSOCIATED BACTERIA IN A BLOOD BANK ENVIRONMENT

**DOI:** 10.64898/2026.07.19.26357333

**Authors:** María Cecilia D’Arpino, Daniel Alonso-Reyes, Mariana Grillo-Puertas, Fátima Silvina Galván, Natalia Noelia Alvarado, Luciano José Martínez, María Gabriela Marranzino, Virginia Helena Albarracín

## Abstract

Blood banks represent highly controlled healthcare environments where microbiological surveillance has traditionally focused on blood products rather than environmental microbial reservoirs. Despite their critical role in transfusion safety, the ecology of surface-associated microorganisms and the persistence traits that enable their long-term survival remain poorly understood. Here, we combined scanning electron microscopy, culture-based microbiology, phenotypic characterization, MALDI-TOF mass spectrometry, and whole-genome sequencing to investigate whether surfaces within a public blood bank facility constitute reservoirs of environmentally derived bacteria with enhanced persistence potential. Samples collected from a public blood bank in Tucumán, Argentina yielded 37 culturable bacterial isolates, predominantly Gram-positive taxa together with a limited number of opportunistic Gram-negative species. More than 30% of the isolates exhibited multidrug resistance, while several strains displayed strong biofilm formation, amyloid-like fiber production, motility, and hemolytic activity, indicating multiple phenotypic strategies associated with long-term surface persistence. Whole-genome sequencing of six representative isolates confirmed species identity, identified genes related to antimicrobial resistance, adhesion, biofilm formation, stress adaptation, and cytotoxicity, and revealed frequent genotype-phenotype discordance, highlighting the importance of integrating genomic and phenotypic analyses. Notably, one isolate exhibited less than 92% average nucleotide identity with publicly available genomes, suggesting the presence of a previously undescribed environmental species. Thus, blood bank surfaces function as selective ecological niches favoring bacteria with persistence-associated traits rather than simply reflecting contamination from blood products. These microorganisms may constitute latent biosafety hazards if sanitization barriers fail, particularly in facilities handling biological materials intended for vulnerable patients. Our results support the incorporation of integrated bioimaging, phenotypic characterization, and genome-resolved environmental surveillance into infection prevention strategies and transfusion biosafety programs within a One Health framework.

## INTRODUCTION

Healthcare environments are increasingly recognized as ecological niches where diverse microbial communities can persist and spread, despite routine cleaning and disinfection procedures (Dancer, 2009; Dancer, 2023). While direct patient-to-patient transmission remains a major factor in healthcare-associated infections (HAIs), growing evidence shows that inanimate surfaces—such as countertops, medical equipment, and air conditioning systems—also serve as reservoirs for opportunistic and pathogenic microorganisms (Weber et al., 2013; Suleyman et al., 2018). These surfaces act as contact points for healthcare personnel and patients, facilitating cross-contamination and microbial persistence under suboptimal environmental conditions (Facciolà et al., 2019). The long-term survival of potential pathogens in clinical settings has been linked to outbreaks and infection clusters, emphasizing the need for continuous monitoring and more effective control strategies (Otter et al., 2013).

One of the most pressing challenges in these settings is the presence of bacteria exhibiting multiple virulence traits that support their survival, dissemination, and disease-causing potential (Mühlen and Dersch, 2016). Among the most studied are biofilm formation, motility, hemolytic activity, and amyloid-like fiber production. These features allow bacteria to firmly adhere to surfaces, resist disinfectants, evade host immune responses, and even cause direct cellular damage (Campos et al., 2020). Biofilms represent structured communities embedded in a self-produced extracellular matrix composed of polysaccharides, proteins, and extracellular DNA, which form a physical and chemical barrier against antimicrobial agents. These matrices enhance interspecies cooperation, promote horizontal gene transfer, and are strongly associated with persistent colonization in clinical settings (Costerton et al., 1995; Flemming and Wingender, 2010; Lazăr and Chifiriuc, 2010; Johnson and Russo, 2002).

Antimicrobial resistance (AMR) further complicates this scenario, especially in healthcare settings where extensive antibiotic use creates selective pressure for the emergence of multidrug-resistant (MDR), extensively drug-resistant (XDR), and even pan-drug-resistant (PDR) strains (Magiorakos et al., 2012). Pathogens such as *Staphylococcus aureus*, *Acinetobacter baumannii*, and *Pseudomonas aeruginosa* are frequently implicated in difficult-to-treat HAIs due to the co-occurrence of resistance genes and virulence factors. This convergence severely limits treatment options and contributes to prolonged hospital stays, higher mortality, and increased healthcare costs (Miller and Arias, 2024). Effective prevention thus requires not only new therapeutic strategies but also the implementation of environmental surveillance programs and molecular tools to identify and monitor high-risk strains (Miller and Arias, 2024; Liu et al., 2026).

Despite extensive research on microbial contamination in surgical areas, intensive care units, and emergency departments, (Totaro et al., 2019; Ribeiro et al., 2019; Osman et al., 2024), blood banks—facilities essential for transfusion services—have been largely overlooked in environmental microbiome studies. These units handle and store biological materials under conditions that, while carefully regulated, may still support microbial persistence. In particular, the manipulation of blood products, temperature-controlled storage, and the use of automated processing systems create microenvironments that are potentially favorable to bacterial survival and adaptation. Given that transfusion recipients are often immunocompromised, understanding the microbial landscape of these environments is crucial to minimizing the risk of contamination and ensuring biosafety throughout the transfusion chain (Quaranta et al., 2015; Shouman et al., 2023).

In this context, the present study aimed to characterize for the first time the microbiome associated with the surfaces of a public blood bank facility in Tucumán, Argentina. Specifically, we investigated bacterial isolates recovered from critical areas within the facility—namely the production, distribution, and molecular biology laboratories—using a combination of bioimaging, phenotypic assays and whole-genome sequencing. Our objectives were to determine the taxonomic identity of the isolates, assess antimicrobial resistance profiles and virulence-linked traits (biofilm formation, motility, hemolytic activity, and curli-like fiber production), and explore the genetic basis of these features.

## MATERIALS AND METHODS

Sampling site and procedures. This study was conducted at the Central Blood Bank “Dr. César Guerra,” which operates under the Integrated Health Program (PRIS-SI.PRO.SA: Programa de Salud Integral - Sistema Provincial de Salud) of Tucumán Province, Argentina. The Blood Bank is a critical public health facility responsible for the collection, processing, storage, and distribution of blood products. Whole blood obtained from donors is processed in the Production and Fractionation Department to prepare various components, including erythrocyte concentrates, platelet concentrates, fresh frozen plasma, cryoprecipitate-reduced plasma, and cryoprecipitate. Processing involves refrigerated centrifugation at 19 °C using Presvac DP-2065 R Plus centrifuges of concentrates and their subsequent maintenance under continuous agitation using Presvac AP-48 L platelet shakers at 18–22 °C. The Molecular Biology Department for Transfusion-Transmitted Infections (TTI) is tasked with nucleic acid screening for human immunodeficiency virus (HIV), human hepatitis C (HCV), and human hepatitis B (HBV) using an automated system that includes a Hamilton MICROLAB STAR IVD/STARlet IVD pipettor, COBAS® AmpliPrep for PCR preparation, and COBAS® TaqMan® Analyser for real-time PCR detection. Sampling was carried out in December 2019, following established biosafety guidelines. Surface swabs were taken from various areas in the Production, Distribution, and Molecular Biology departments. Specific sampling sites included air conditioners, countertops, centrifuges, reagent tables, and specialized equipment such as the platelet agitator and COBAS systems. The sampling sites were coded as follows: Production - air conditioner (AA-PRO), worktables (ME-PRO), centrifuge (E-PRO), Distribution - platelet agitator (AP-DIS); Molecular Biology - air conditioner (AA-BM), reagent tables (MR-BM), other worktables (ME-BM), and COBAS equipment (E-BM).

For the direct observation of microorganisms on the sampling sites surfaces, freshly-open 3M™ adhesive tape was used as described previously (Alonso-Reyes et al., 2025). Strips of 2–3 cm were cut in advance using sterile scissors under aseptic conditions and sterilized by exposure to germicidal UV-C radiation for 15 minutes. Subsequently, the strips were gently adhered to the different surfaces and then transferred to 2 ml tubes containing Karnovsky’s fixative solution (2.66% w/v paraformaldehyde and 1.66% w/v glutaraldehyde in 0.1 M phosphate buffer, pH 7.2), for transportation to the lab.

Bacterial isolation and identification. Samples were plated onto sterile Petri dishes containing Luria-Bertani (LB) agar medium (1% tryptone, 1% NaCl, 0.5% yeast extract, 1.5% agar, pH 7.0) supplemented with cycloheximide (10 μg mL−1) to inhibit fungal growth. Each swab was streaked on duplicate plates and incubated at 30 °C for 24–48 hours. Half of the plates were supplemented with nalidixic acid (10 μg mL−1) to prevent the growth of fast-growing Gram-negative bacteria (GNB). Colonies were subcultured individually onto fresh LB agar plates for purification and maintained through successive streaking. Preliminary identification involved morphological, microscopic, and phenotypic analyses (Cheesbrough, 2006). Morphological characterization was performed using Gram staining, light and scanning electron microscopy. For SEM, cell cultures grown in LB broth at 30 °C and 180 rpm until the exponential phase were centrifuged, washed in 0.9% NaCl, and fixed in Karnovsky’s solution. For molecular identification, isolates were analysed using the VITEK MS system (bioMérieux), a MALDI-TOF-based platform. All strains in axenic forms were kept in lyophilised form at room temperature and as glycerol stocks in refrigerated conditions (-20 °C). They were then incorporated into our Culture Collection called Cepario de Bacterias Ambientales del CIME (CEBaC), within the Catalogue Urban Bacteria, BS division (Table 1).

**Table 1.**
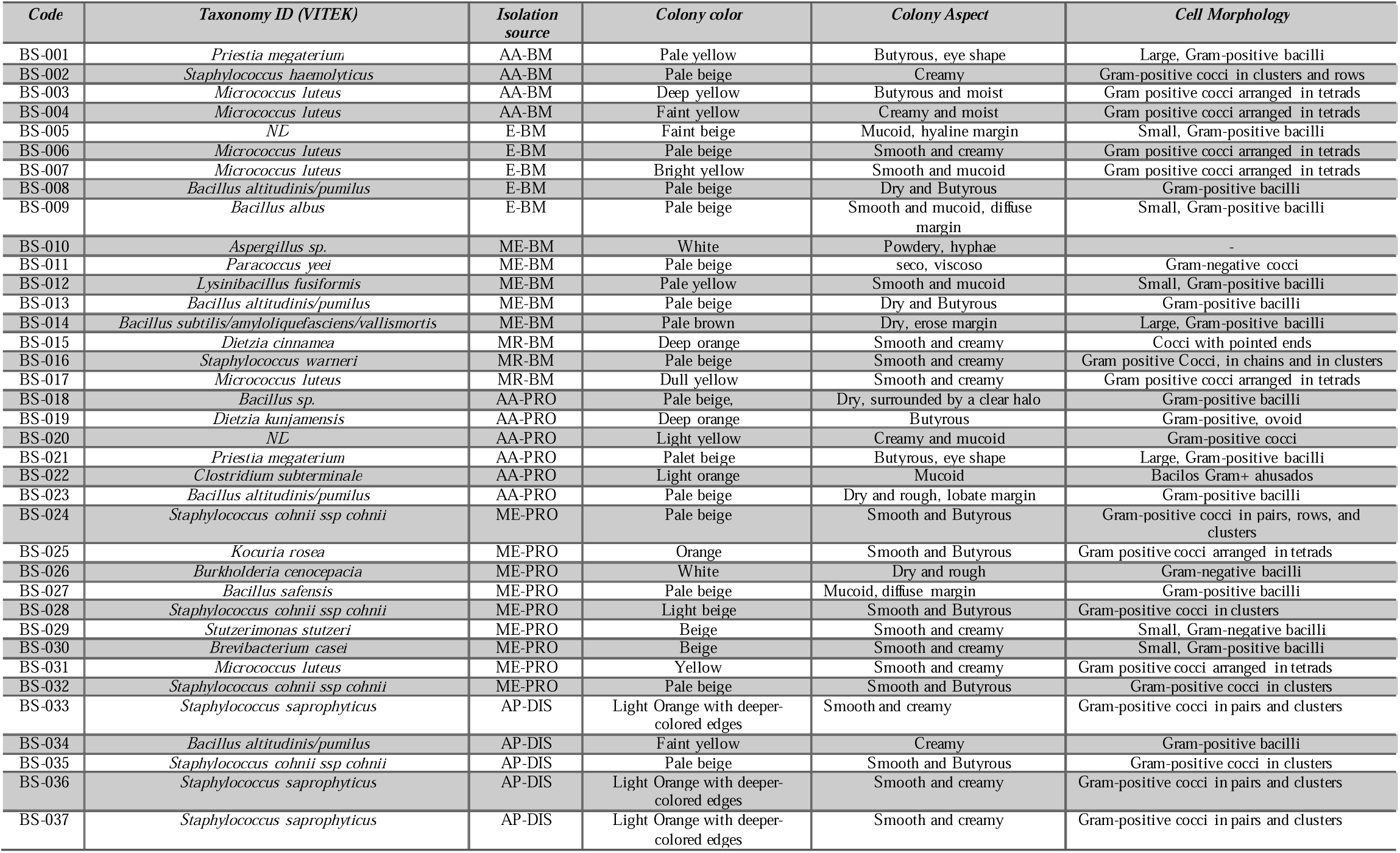
Culturable microorganisms recovered from environmental surfaces of the Central Blood Bank “Dr. Cesar Guerra” in Tucuman, Argentina. The table summarizes the 37 isolates obtained from surface swabs collected in the Molecular Biology, Production, and Distribution areas and recovered on LB agar at 30 degrees C. Isolates are listed by institutional BS code and include taxonomic identification by VITEK MS, isolation source, colony pigmentation and appearance, and cell morphology determined by Gram staining and light microscopy. The strain collection was incorporated into the CIME Environmental Bacteria Collection (CEBaC), Urban Bacteria Catalogue, BS division. Sampling-site codes are: AA-BM, Molecular Biology air conditioner; MR-BM, Molecular Biology reagent tables; ME-BM, Molecular Biology worktables; E-BM, Molecular Biology COBAS equipment; AA-PRO, Production air conditioner; ME-PRO, Production worktables; E-PRO, Production centrifuge; AP-DIS, Distribution platelet agitator. ND, not determined.

Scanning Electron Microscopy (SEM). Tape-strips with samples of blood bank surfaces and isolated strains were processed for SEM imaging as described previously (D’Arpino et al., 2020; Alonso-Reyes et al., 2025; Galván and Albarracín, 2026). Briefly, fixed samples were dehydrated through ethanol gradients (50, 70, 90 and 100%), treated with acetone, and dried using a critical point dryer (Denton Vacuum DCP-1). They were mounted on stubs with conductive tape, gold-coated with a JEOL JFC-1100 sputter coater and examined with a Zeiss SUPRA 55VP SEM at 3 kV using the SE2 detector at the electron microscopy core facility of UNT-CONICET: Centro Integral de Microscopía Electrónica (CIME).

Antibiotic Susceptibility Testing. Antibiograms were performed using the disk diffusion method on Mueller-Hinton agar (BB-NCIPD Ltd.), following CLSI (2020) and EUCAST guidelines (2026). Antibiotic (ATB) panels were selected based on national and international recommendations. Specific ATBs tested varied by bacterial group (Table 2). Inocula were prepared to a 0.5 McFarland standard and antibiotic disks were placed on seeded plates. After 24 h at 37 °C, inhibition zones were measured and interpreted. Multidrug resistance (MDR) was defined as resistance to ≥1 agent in ≥3 antimicrobial classes (Magiorakos et al., 2012).

**Table 2.**
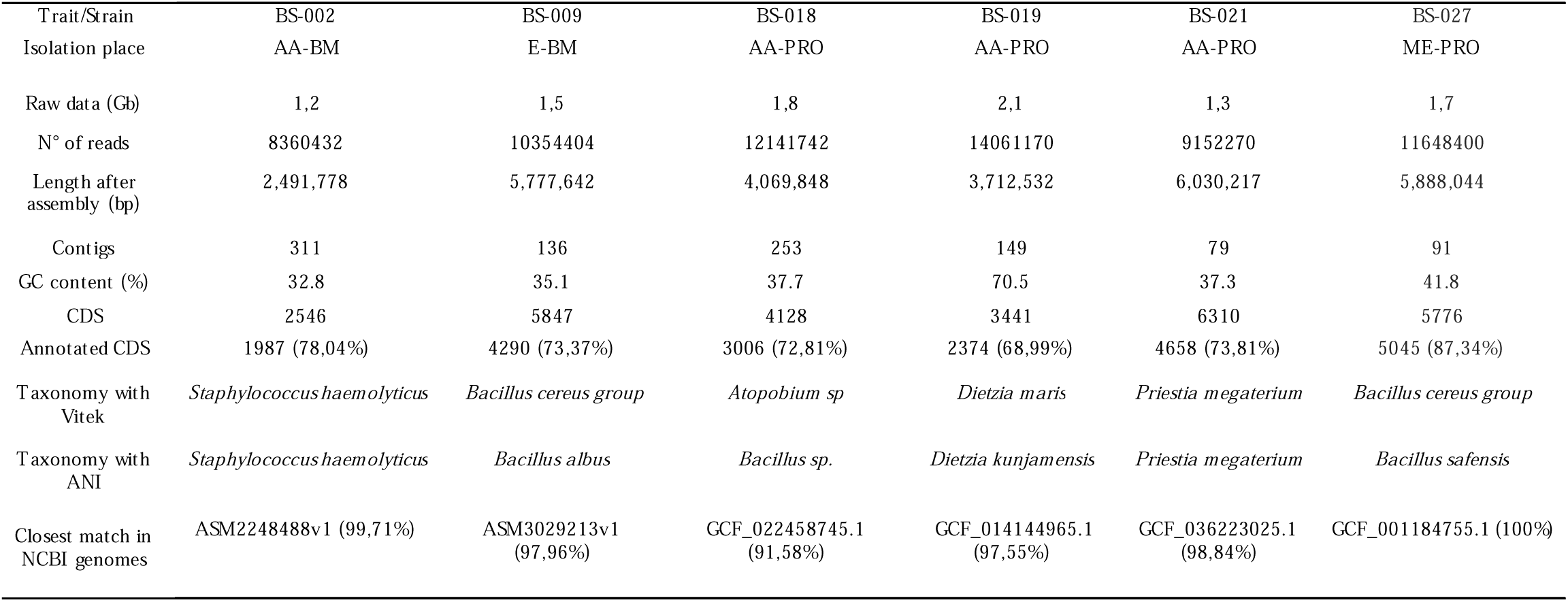
Genome sequencing, assembly, annotation, and sequence-based taxonomic assignment of six representative blood-bank surface isolates. Whole-genome sequencing was performed for strains BS-002, BS-009, BS-018, BS-019, BS-021, and BS-027 using Illumina NovaSeq PE150 reads. The table reports isolation site, raw sequence yield, read number, de novo assembly size, contig number, GC content, predicted coding sequences (CDS), annotated CDS, VITEK MS taxonomic assignment, average nucleotide identity (ANI)-based assignment, and the closest available NCBI genome match. Assemblies were generated with SPAdes v3.15.4, taxonomic classification was refined using Kraken2 and ANI comparisons, and functional annotation was performed with the Arche pipeline v1.0.1. ANI confirmed species-level identity for five genomes, whereas BS-018 showed less than 92% similarity to publicly available genomes, below the commonly used >=95% species boundary, suggesting that this isolate may represent a novel environmental taxon. Abbreviations: Gb, gigabases; bp, base pairs; GC, guanine-cytosine content; CDS, coding DNA sequences; ANI, average nucleotide identity.

Biofilm formation assay and colony morphotypes. Biofilm production was evaluated in 96-well polystyrene plates using a crystal violet staining method (O’Toole and Kolter, 1998). After incubation at 30 °C, wells were washed, stained with 0.1% crystal violet, and absorbance was measured at 595 nm. Biofilm formation capacity was classified following Stepanović et al. (2007): non-producers (-), weak (+), moderate (++), strong (+++), or robust (++++) based on comparison with blank controls. Curli and amyloid-like fiber production were evaluated on LB agar supplemented with Congo Red (40 µg/mL) and Brilliant Blue (20 µg/mL). Colony color and texture were assessed after 96 h at 30 °C (Da Re and Ghigo, 2006).

Motility and hemolytic activity assays. Motility was assessed on 0.3% semi-solid LB agar. Inoculated plates were incubated at 30 °C, and colony spread was measured. Colonies with a diameter of 0.5–1 cm were considered motile (+), and those >1 cm were denoted as highly motile (++). Hemolysis was tested on blood agar. After 24 h at 30 °C, colonies were inspected for clear (beta), greenish (alpha), or absent (gamma) hemolysis zones (Buxton, 2005).

DNA Extraction, Whole Genome Sequencing (WGS) and bioinformatic analysis. Six strains from different isolation sources and differential phenotype were selected for WGS (Table 2). DNA was extracted using DNA Puriprep B-kit (Inbio Highway, Argentina). Gram-positive cells were pretreated with lysozyme; all samples underwent proteinase K digestion, RNase treatment, and ethanol precipitation. DNA was purified using spin columns and eluted in 60 µL buffer. Concentration and quality were assessed with a µDrop plate (Thermo Scientific). Genomes were sequenced by Novogene UK using Illumina NovaSeq PE150. Assemblies were generated with SPAdes v3.15.4. Sequencing generated ∼65 million reads across all samples, averaging ∼10 million reads and 1.6 Gb per genome. The number of contigs ranged from 79 (BS-021) to 311 (BS-002). Taxonomic classification was performed using Kraken2 with the PlusPF database. ANI values were computed using PyANI to confirm species-level identity. Functional annotation was performed using the Arche pipeline v1.0.1 (Alonso-Reyes and Albarracín, 2025). The lowest annotation (69%) was obtained with the genome of strain BS-019, and the highest (∼87%) was in CH-027. Table 2 presents data on the isolates, including their isolation locations, sequencing results, de novo assemblies, and annotations.

## RESULTS

### Environmental bacteria extensively colonize blood bank surfaces

Sampling revealed widespread microbial colonization across the three main operational areas of the blood bank, including the Molecular Biology, Production, and Distribution laboratories. Fourteen of the sixteen sampled plates (87.5%) yielded microbial growth, resulting in the recovery of 37 culturable isolates that were subsequently purified and characterized (Fig. 1; Table 1). The culturable community was dominated by Gram-positive bacteria, which accounted for the vast majority of recovered isolates, whereas only three Gram-negative species were detected. Members of the genera *Micrococcus*, *Bacillus*, *Staphylococcus*, *Dietzia*, *Lysinibacillus*, *Brevibacterium*, *Kocuria*, *Clostridium*, *Paracoccus*, *Burkholderia*, and *Stutzerimonas* were identified by MALDI-TOF MS (Fig. 1). Despite supplementing the isolation medium with cycloheximide to inhibit fungal growth, a fungal isolate (*Aspergillus* sp.) was recovered from a molecular biology bench. This strain was retained in CeMAC but was not included in subsequent analyses.

**Figure 1.**
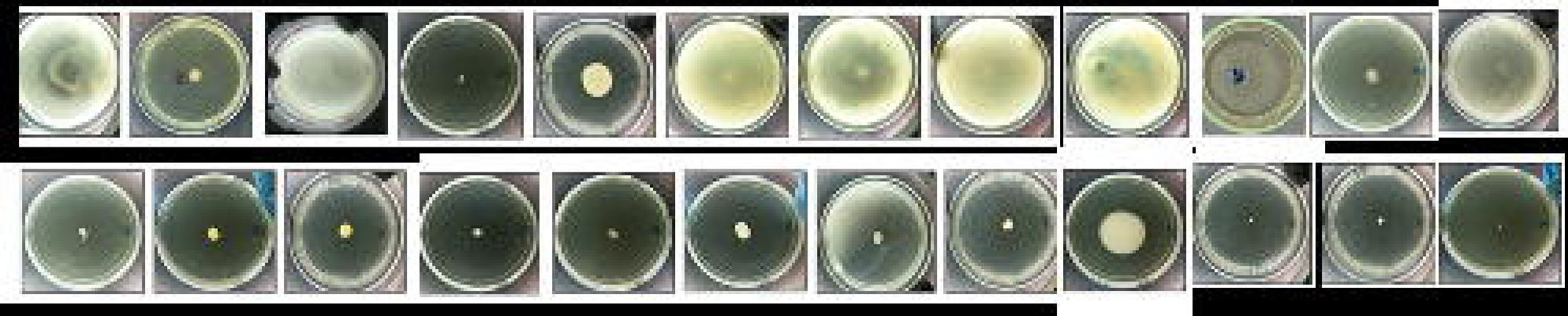
Distribution and taxonomic composition of culturable microorganisms recovered from environmental surfaces of the blood bank. (A) Number of bacterial isolates recovered from the three operational areas of the facility: Molecular Biology, Production, and Distribution. (B) Relative abundance of bacterial genera recovered from all sampled surfaces. (C) Heatmap showing the distribution of bacterial genera across the three operational areas. (D) Bubble plot summarizing the spatial distribution of bacterial genera throughout the blood bank.

Taxonomic composition differed among operational areas. The Molecular Biology laboratory was characterized by the predominance of *Micrococcus luteus*, particularly on air-conditioning systems, workbenches, and laboratory equipment, whereas the Production and Distribution areas exhibited greater taxonomic diversity and included all Gram-negative isolates recovered in this study. Species of *Bacillus*, *Staphylococcus*, and *Burkholderia* were preferentially associated with processing equipment, platelet agitators, and work surfaces directly involved in blood component handling (Table 1). These results indicate that viable bacteria are broadly distributed throughout the blood bank facility, suggesting that multiple operational surfaces constitute stable reservoirs of microorganisms adapted to healthcare-associated indoor environments.

### Bioimaging reveals structured microbial colonization of critical blood bank surfaces

Scanning electron microscopy provided direct evidence that bacterial colonization occurred predominantly as structured surface-associated communities rather than as isolated cells. Biofilms and extracellular polymeric matrix (EPS) were consistently observed on representative surfaces from all operational areas, including workbenches, centrifuges, and platelet agitators (Fig. 2). Cells were frequently embedded within an abundant extracellular matrix or completely encapsulated by EPS, indicating mature biofilm development on routinely used equipment.

**Figure 2.**
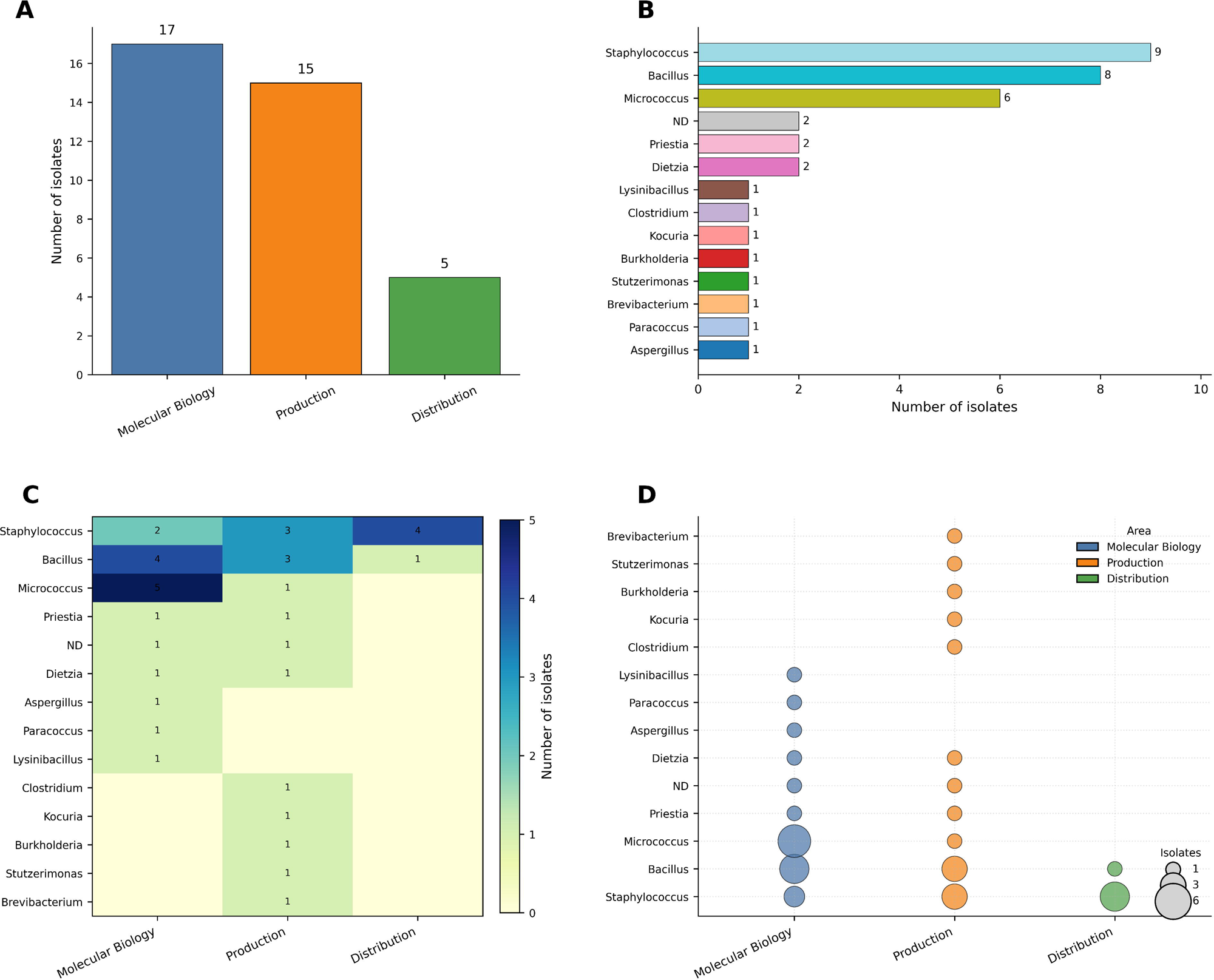
Scanning electron micrographs of tape-strips sampled on the Blood Bank Facility laboratories surfaces. Images were acquired using SEM on a Supra 55 VP instrument at 20.00 kx magnification. WD. 3.9 mm, EHT 2.00 kV, secondary electron detector (SE2). a, BM workbench, b, PRO workbench, c, DIS platelet shaker, d, PRO centrifuge. Arrows indicate cells immersed in extracellular matrix (EPS) components (a, b, and c) or encapsulated in EPS (d).

SEM examination of representative isolates further revealed considerable morphological diversity among recovered bacteria, including coccoid and rod-shaped morphotypes, differences in cell size, surface texture, and colony architecture (Fig. 3). The combination of ultrastructural observations and colony morphology confirmed that the isolated strains represented taxonomically diverse microorganisms capable of colonizing abiotic surfaces under blood bank operating conditions. Together, SEM observations consistently revealed organized microbial colonization associated with extracellular matrix production and biofilm-like structures across multiple blood bank surfaces.

**Figure 3.**
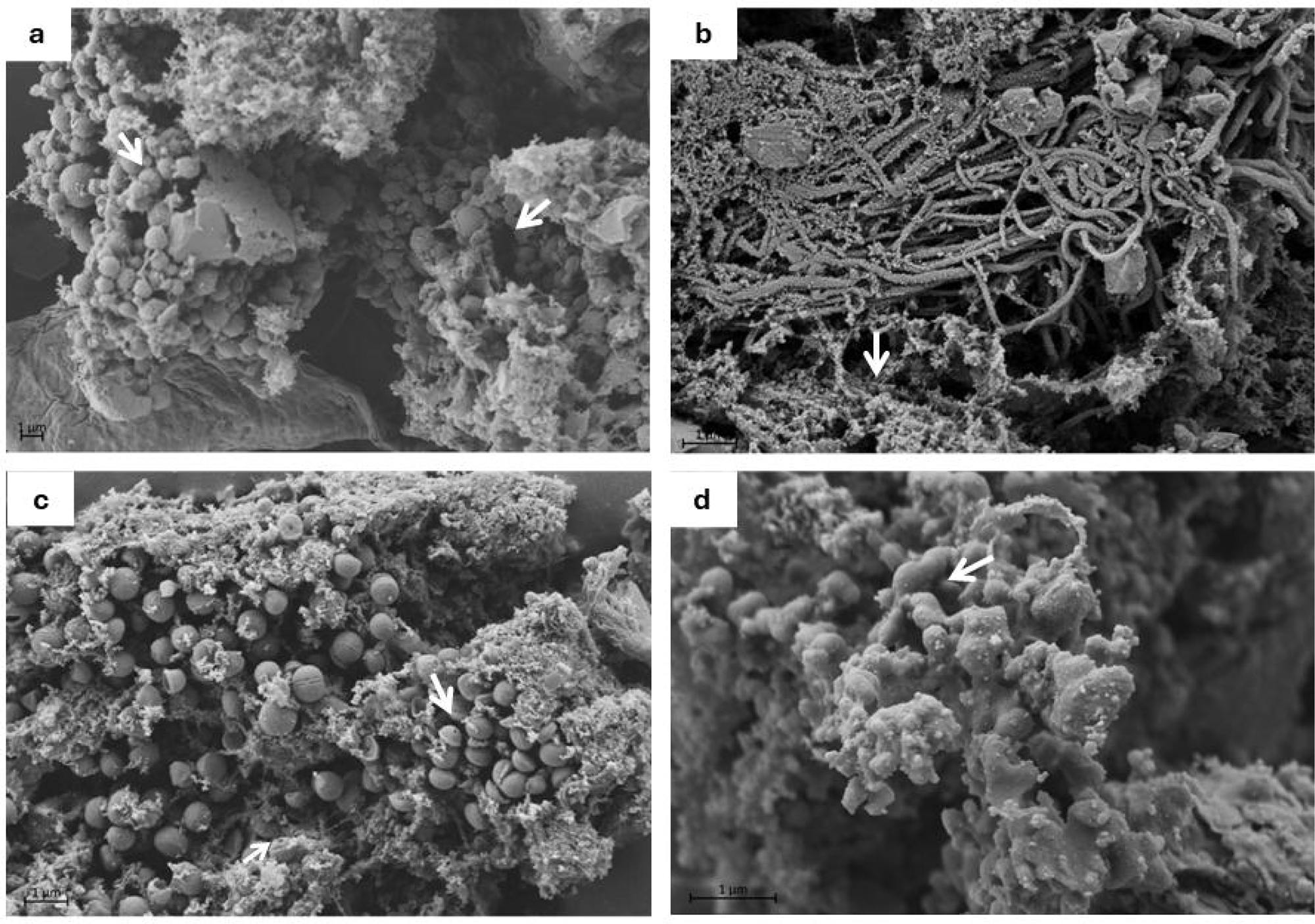
Scanning electron micrographs of BBI. Images were obtained using SEM on a Supra 55 VP instrument at 5.00 kx (bar), a working distance (WD) of 3.9 mm, an EHT of 2.00 kV, and a secondary electron detector (SE2). The insets in each image evidence the phenotypic appearance of each strain in LB medium at pH 7. From the molecular biology laboratory: a, *Bacillus megaterium* (air-conditioned); b, *Bacillus altitudinis*/pumilus (equipment); c, *Bacillus albus* (equipment); d, *Paracoccus yeei* (workbench); e, *Lysinibacillus fusiformis* (workbench); f, *Bacillus subtilis*/amyloliquefaciens/vallismortis (workbench). g, *Dietzia cinnamea* (reagent table). h, *Staphylococcus warneri* (reagent table). From the production laboratory, i, *Micrococcus luteus* (air conditioning); j, *Staphylococcus cohnii* ssp. cohnii (banks); k, *Burkholderia cenocepacia* (banks); l, *Bacillus safensis* (banks); m, *Brevibacterium casei* (banks); n, *Staphylococcus saprophyticus* (platelet shaker); o, *Priestia megaterium* (air conditioning). Images are representative of at least three independent observations.

### Blood bank isolates (BBI) display multiple persistence-associated phenotypes

Phenotypic characterization revealed that many BBI expressed complementary traits associated with long-term persistence on abiotic surfaces. More than 30% of the recovered bacteria fulfilled the criteria for multidrug resistance (MDR), while several isolates simultaneously exhibited biofilm formation, extracellular matrix production, motility, and hemolytic activity, indicating that persistence within the blood bank environment relies on multiple adaptive strategies rather than on a single virulence-associated characteristic (Table 3, 4 and 5). These phenotypes were distributed across phylogenetically diverse taxa, suggesting that persistence represents a convergent ecological strategy shared by unrelated environmental bacteria.

**Table 3.**
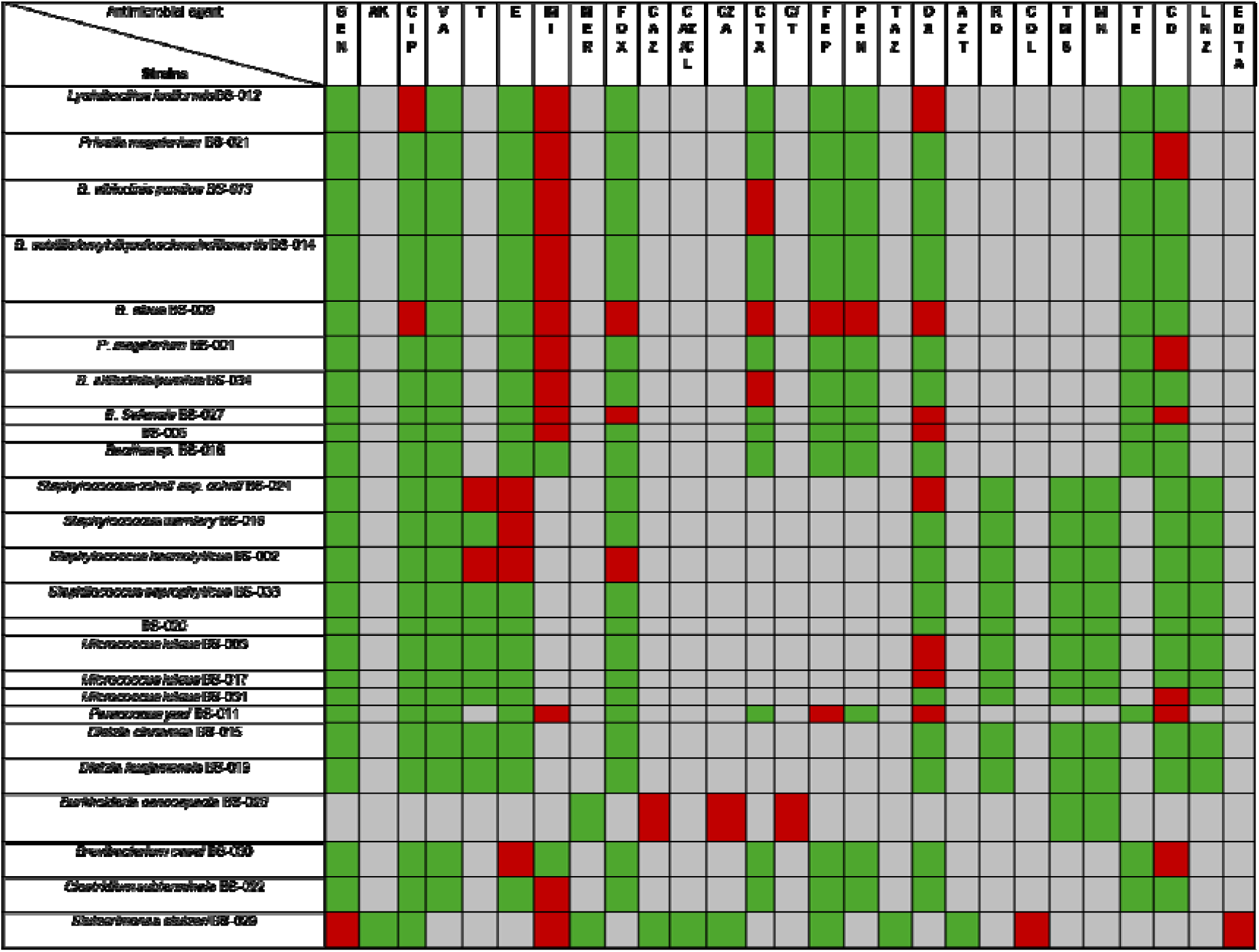
Antibiotic susceptibility profiles of all test organisms as determined by disc diffusion assays. The table shows representative strains isolated from the molecular biology (MB), production (PRO) and distribution (DIS) laboratories of the blood bank. The different groups of microorganisms received a specific antimicrobial assignment as detailed below. Antibiograms for Gram-positive bacilli (MBL). GEN (Gentamicin 120 µg), OX (Oxacillin 1 µg), CIP (Ciprofloxacin 5 µg), VA (Vancomycin 30 µg), E (Erythromycin 15 µg), TE (Tetracycline, 30 µg), IMI (Imipenem 10 µg), FOX (Cefoxitin 30 µg), CD (Clindamycin 2 µg) CTX (Cefotaxime 30 µg), CEF (Cefepime 30 µg), PEN (Penicillin 10 µg). Antibiograms for *Staphylococcus*. GEN (Gentamicin 120 µg), RD (Rifampicin 5 µg), OX (Oxacillin 1 µg), CIP (Ciprofloxacin 5 µg), VA (Vancomycin 30 µg), CD (Clindamycin 2 µg), E (Erythromycin 15 µg), LNZ (Linezolid 30 µg), MN (Minocycline 30 µg), FOX (Cefoxitin 30 µg), TMS (trimethoprim-sulfamethoxazole 25 µg), T (Teicoplanin 30 µg), CLI (Clindamycin 2 µg). Antibiograms for *Stutzerimonas stutzeri*. CAZ (Ceftazidime 30 µg), CAZ/CL (Ceftazidime + clavulanic acid 30/10 µg), FEP (Cefepime 30 µg), TAZ (Piperacillin + tazobactam 100/10 µg), EDTA, IMI (Imipenem 10 µg), CZA (Ceftazidime-avibactam 14 µg), COL (Colistin 10 µg), AZT (Aztreonam), GEN (Gentamicin 120 µg), AK (Amikacin 30 µg), CIP (Ciprofloxacin 5 µg). Antibiograms for *Burkholderia cenocepacia*. MRP (Meropenem 10 µg), CZA (Ceftazidime-avibactam 14 µg), TMS (trimethoprim-sulfamethoxazole 25 µg), MN (Minocycline 30 µg), CAZ (Ceftazidime 30 µg), C/T (Ceftolozane/tazobactam 40 µg). Resistant (R; Red); Susceptible (S; Green); NR, natural resistance, antibiotics not tested (gray).

Among Gram-positive bacilli, ATB resistance was most frequently associated with carbapenems, particularly imipenem, whereas susceptibility to gentamicin, vancomycin, erythromycin, and tetracycline was generally preserved. Staphylococcal isolates displayed variable resistance patterns, including resistance to macrolides, β-lactams, glycopeptides, and aminoglycosides, with *Staphylococcus haemolyticus* and *Staphylococcus cohnii* exhibiting MDR phenotypes. In contrast, all recovered Gram-negative isolates (*Burkholderia cenocepacia*, *Stutzerimonas stutzeri*, and *Paracoccus yeei*) fulfilled the criteria for multidrug resistance, although each species displayed a distinct resistance profile. Interestingly, *Bacillus albus* BS-009 exhibited the broadest resistance spectrum among Gram-positive bacilli, whereas *Staphylococcus haemolyticus* BS-002 combined methicillin resistance with a multidrug-resistant phenotype. Likewise, all three Gram-negative isolates showed resistance to multiple antimicrobial classes, reinforcing their potential relevance as opportunistic environmental reservoirs of antimicrobial resistance.

Rather than being restricted to a particular taxonomic group, multidrug resistance occurred in strains displaying diverse ecological and phenotypic characteristics, suggesting that antimicrobial resistance constitutes one component of a broader persistence strategy. Importantly, MDR isolates frequently co-occurred with additional persistence-associated traits, including biofilm formation and motility, reinforcing their potential to survive repeated cleaning and environmental stress. Indeed, biofilm formation was widely distributed among the environmental isolates, although the intensity of biofilm production varied substantially between strains. Strong biofilm producers were identified in several genera, including *Bacillus*, *Dietzia*, *Burkholderia*, and *Staphylococcus*, whereas other isolates exhibited only weak or undetectable biofilm formation under the experimental conditions tested. Interestingly, strong biofilm production was observed across phylogenetically unrelated taxa, indicating that surface persistence represents a convergent adaptive strategy rather than a taxon-specific characteristic.

In accordance, colony morphology on Congo Red/Brilliant Blue agar revealed substantial phenotypic diversity among the isolates (Fig. 4, Table 4). Numerous strains developed dry, rough, red or brown colonies with distinct halo patterns, displaying morphotypes previously described as being consistent with the production of extracellular matrix components, including amyloid-like fibers and/or cellulose (Romling et al., 2000; Coleri et al., 2017). These phenotypes were particularly frequent among members of the *Bacillus* and *Staphylococcus* genera, although marked variability was also observed within individual genera. In contrast, other isolates produced smooth or lightly pigmented colonies lacking these characteristic features, suggesting limited or undetectable matrix-associated phenotypes under the experimental conditions evaluated. The widespread occurrence of Congo Red-positive colony morphotypes indicates that the ability to produce extracellular matrix components is common among bacteria colonizing blood bank surfaces. These structures are known to promote surface adhesion, biofilm maturation, and increased tolerance to environmental stresses, thereby contributing to long-term persistence on abiotic surfaces. Nevertheless, because colony morphology on Congo Red agar provides only an indirect indication of extracellular matrix production, these phenotypes should be interpreted as presumptive evidence rather than direct confirmation of amyloid fiber or cellulose biosynthesis.

**Figure 4.**
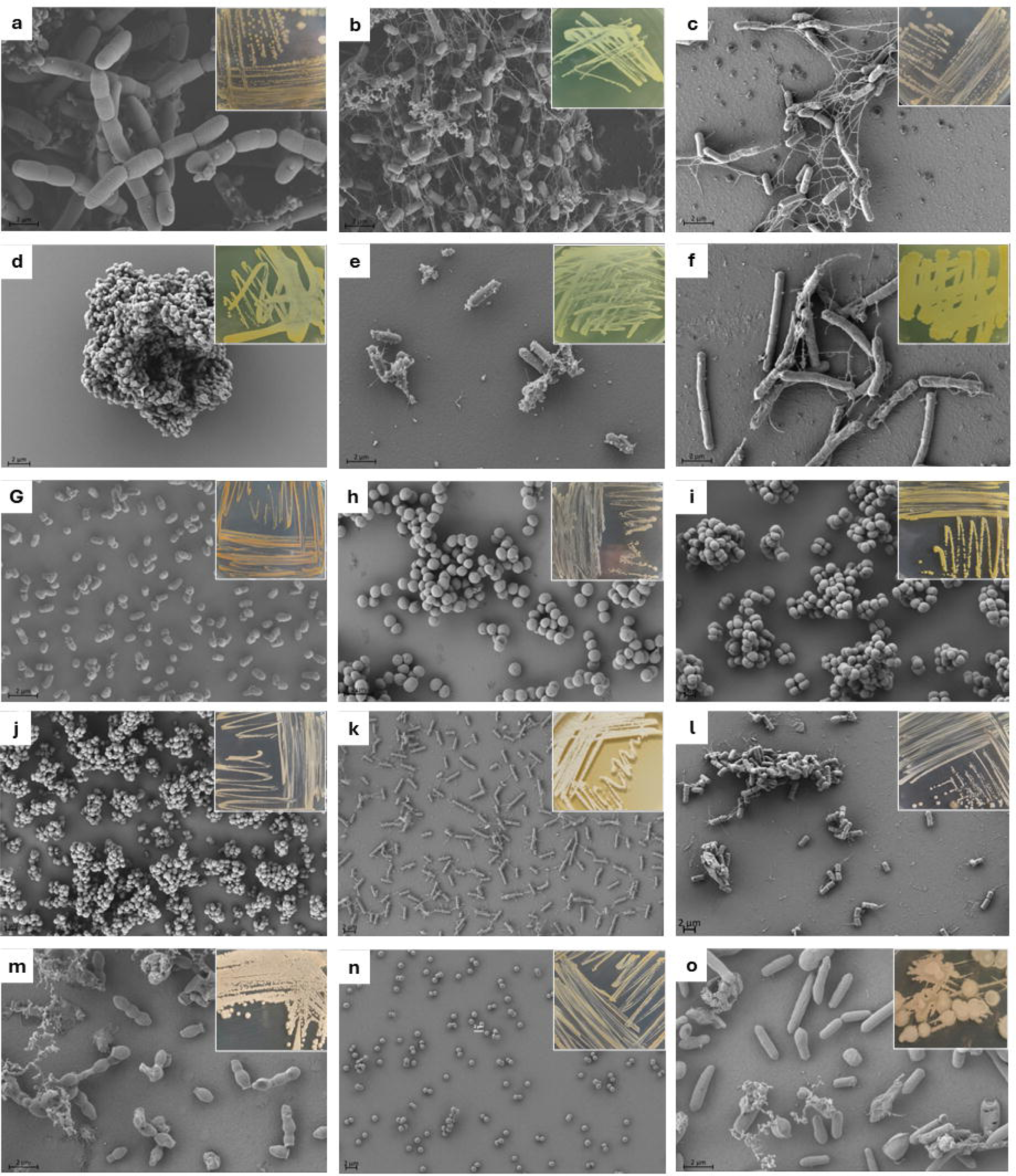
Colony morphotypes of representative BBI grown on LB-agar (low sodium) supplemented with Congo Red and Brilliant Blue dyes. Panels correspond from left to right: Row 1. *Lysinibacillus* sp. BS012; *Bacillus* sp. BS013; *Bacillus* sp. BS014; *Bacillus albus* BS009; *Priestia megaterium*. BS001; *Bacillus* sp. BS018; *Bacillus* sp. BS034; *Bacillus safensis* BS027; *Bacillus* sp. BS005. Row 2: *Priestia megaterium* BS021; *Staphylococcus* sp. BS024; *Staphylococcus* sp. BS016; *Staphylococcus haemolyticus* BS002; *Staphylococcus* sp. BS033; *Staphylococcus* sp. BS020; *Micrococcus* sp. BS003; *Micrococcus* sp. BS017; *Micrococcus* sp. BS031; *Paracoccus* sp. BS011; *Dietzia* sp. BS015; *Dietzia kunjamensis* BS019; *Burkholderia* sp. BS026; Row 3: *Clostridium* sp. BS022; *Brevibacterium* sp. BS030. Images correspond to colonies grown under identical culture conditions. The original scale bar is shown in the lower right corner.

**Figure 5.**
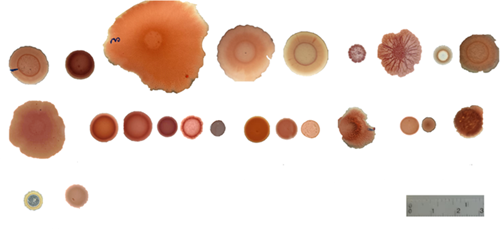
Representative motility phenotypes of BBI. . Images correspond to colonies grown under identical culture conditions. were inoculated onto M63 semi-solid agar (0.3%) using the stab inoculation method. Motility is evidenced by the radial expansion of bacterial growth from the inoculation site after 72 h of incubation. Images are representative of two independent experiments. Panels correspond from left to right: Row 1. *Bacillus* sp. BS005. *Bacillus safensis* BS027, *Bacillus* sp. BS034; *Bacillus* sp. BS018, *Priestia megaterium.* BS001, *Bacillus albus* BS009; *Bacillus* sp. BS014; *Lysinibacillus* sp. BS012; *Bacillus* sp. BS013; *Clostridium* sp. BS022; *Brevibacterium* sp. BS030, *Dietzia kunjamensis* BS019. Row 2: *Micrococcus* sp. BS031;; *Micrococcus* sp. BS017; *Micrococcus* sp. BS003; *Staphylococcus* sp. BS020; *Staphylococcus* sp. BS033; *Staphylococcus haemolyticus* BS002; *Staphylococcus* sp. BS016; *Staphylococcus* sp. BS024; *Priestia megaterium* BS021; *Burkholderia* sp. BS026; *Paracoccus* sp. BS011; *Dietzia* sp. BS015.

**Table 4.**
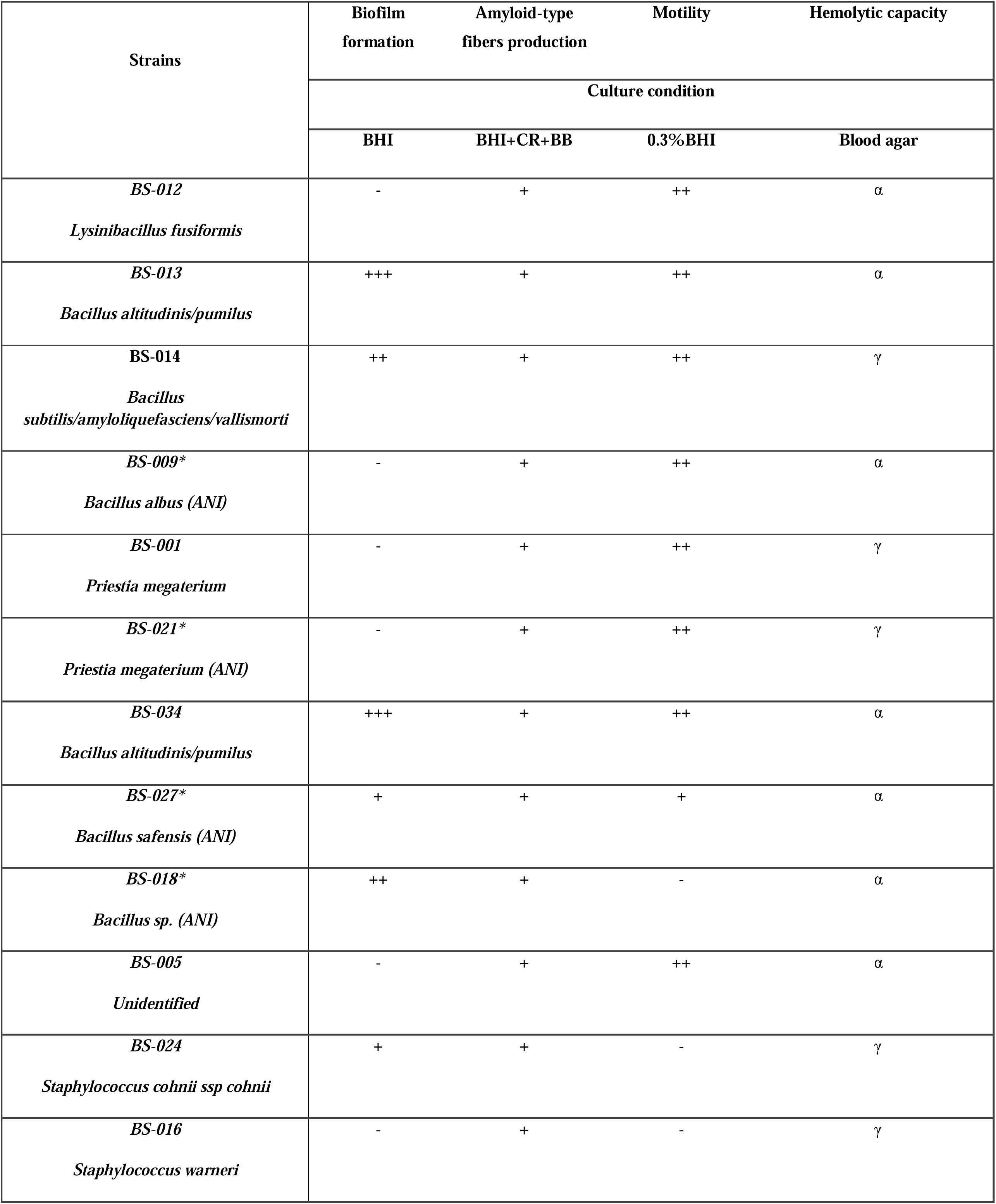
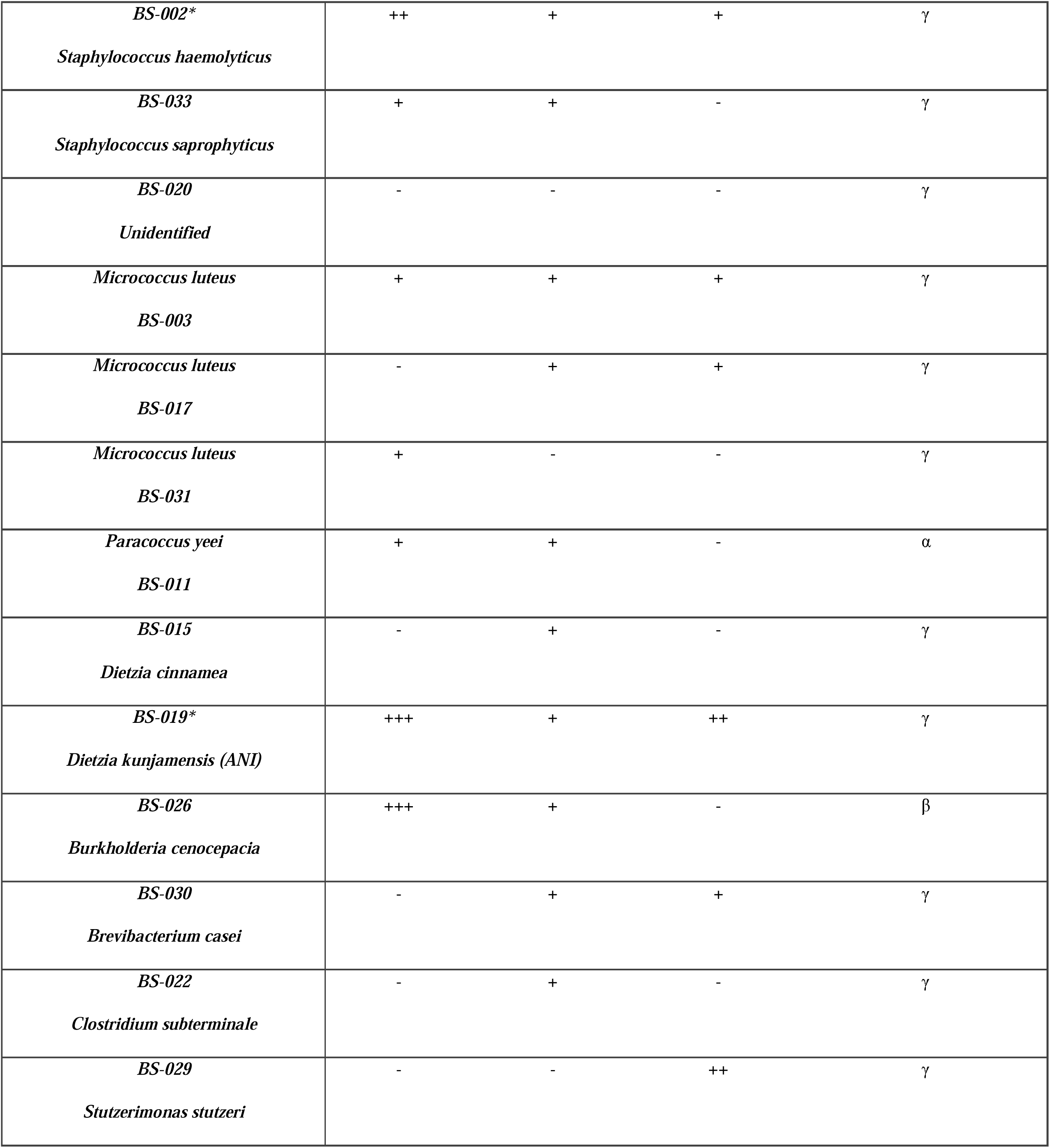
Virulence associated-phenotypes in selected BS strains. Phenotypes were represented as follow: **Biofilm formation:** -, non-biofilm producers; **+**, weak biofilm producers; ++, moderate biofilm producers; +++, strong biofilm producers; and ++++, robust biofilm producers. **Amyloid-like fiber:** +, presence; -, absence. **Motility: -**, no motile; +, colony diameter between 0.5 and 1 cm; and ++, colony diameter more than 1 cm. **Hemolytic capacity:** α, partial lysis; β, complete lysis; γ, no hemolysis. ***\**** Strains analyzed by ANI.

### Genome sequencing reveals strain-specific persistence strategies

Whole-genome sequencing of six representative isolates was performed to investigate the molecular basis underlying the persistence-associated phenotypes observed among blood bank bacteria. The selected strains represented different operational areas and distinct phenotypic profiles, including variations in biofilm formation, antimicrobial resistance, motility, and hemolytic activity. Genome-resolved analyses refined taxonomic identification while providing comprehensive information on the functional repertoire potentially involved in environmental persistence (Table 2).

Genome assemblies ranged from approximately 2.5 to 6.0 Mb and contained between 2,546 and 6,310 predicted coding sequences, with annotation rates ranging from 69% to 87%. Average nucleotide identity (ANI) analyses confirmed species-level assignments for five isolates, including *Staphylococcus haemolyticus* BS-002, *Bacillus albus* BS-009, *Dietzia kunjamensis* BS-019, *Priestia megaterium* BS-021, and *Bacillus safensis* BS-027 (Gupta et al., 2020). In contrast, strain BS-018 shared only 91.58% nucleotide identity with its closest genome available in public databases, well below the accepted species threshold of approximately 95% (Richter and Rosselló-Móra, 2009), suggesting that this isolate may represent a previously un-described healthcare-associated taxon. Pairwise genome comparisons against public databases revealed that the closest relatives of BBI were predominantly isolated from environmental sources; BS-021 showed 98.84% ANI with *Priestia megaterium* BD-304 isolated from bear feces, while BS-027 matched with *Bacillus* sp. FJAT-27238 (100% ANI), a soil bacterium (Liu et al., 2015). Similarly, BS-009 exhibited 97.96% ANI with *Bacillus albus* SXL388 from Chinese soil (Song et al., 2024), and BS-019 matched with *Dietzia kunjamensis* K30-10T (97.55% ANI), isolated from cold desert soils of the Indian Himalayas (Fang et al., 2021). BS-002 was the only strain closely related to an opportunistic pathogen strain from the clinical setting; it shared 99.71% ANI with the *Staphylococcus haemolyticus* 092P2FS1 strain isolated from a bronchoalveolar lavage sample from a hospital in China (Mingyang et al., 2022). These findings illustrates the diversity of surface-associated microorganisms inhabiting blood bank surfaces and highlights the value of genome sequencing for refining taxonomic assignments beyond routine MALDI-TOF identification. An integrated overview of the taxonomic, phenotypic, ultrastructural, and genomic characteristics of the six representative isolates is summarized in Table 5.

**Table 5.**
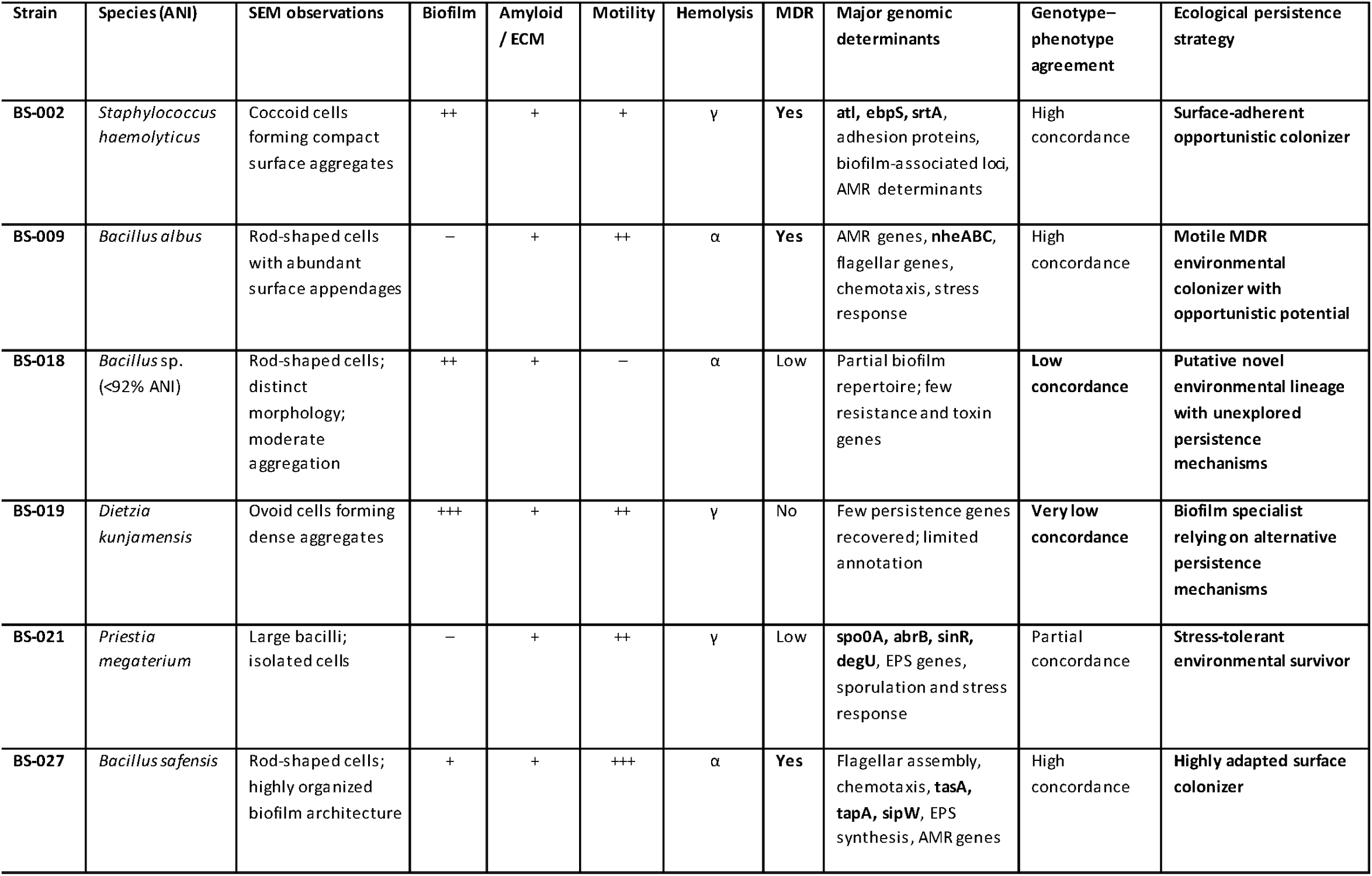

In addition, functional annotation identified a broad repertoire of genes associated with bacterial persistence, including determinants involved in biofilm formation, surface adhesion, extracellular matrix production, antimicrobial resistance, stress adaptation, cytotoxicity, and motility (Fig. 6). The abundance and distribution of these functional categories differed markedly among isolates, indicating that persistence is achieved through strain-specific adaptive strategies rather than through a conserved genomic profile. In general, biofilm-related functions predominated across all the analyzed genomes, whereas hemolysis/cytotoxicity-associated annotations were comparatively restricted (Fig. 6-8). Among the sequenced genomes, *Bacillus safensis* BS-027 and *Bacillus albus* BS-009 exhibited the largest repertoire of persistence-associated genes. Both genomes contained numerous annotations related to motility, antimicrobial resistance, extracellular matrix production, and biofilm-associated functions, suggesting a broad adaptive capacity for colonization and survival on abiotic surfaces. In contrast, *Dietzia kunjamensis* BS-019 contained comparatively fewer annotated persistence determinants despite exhibiting a robust biofilm phenotype, indicating that alternative or poorly characterized mechanisms may contribute to its persistence.

**Figure 6.**
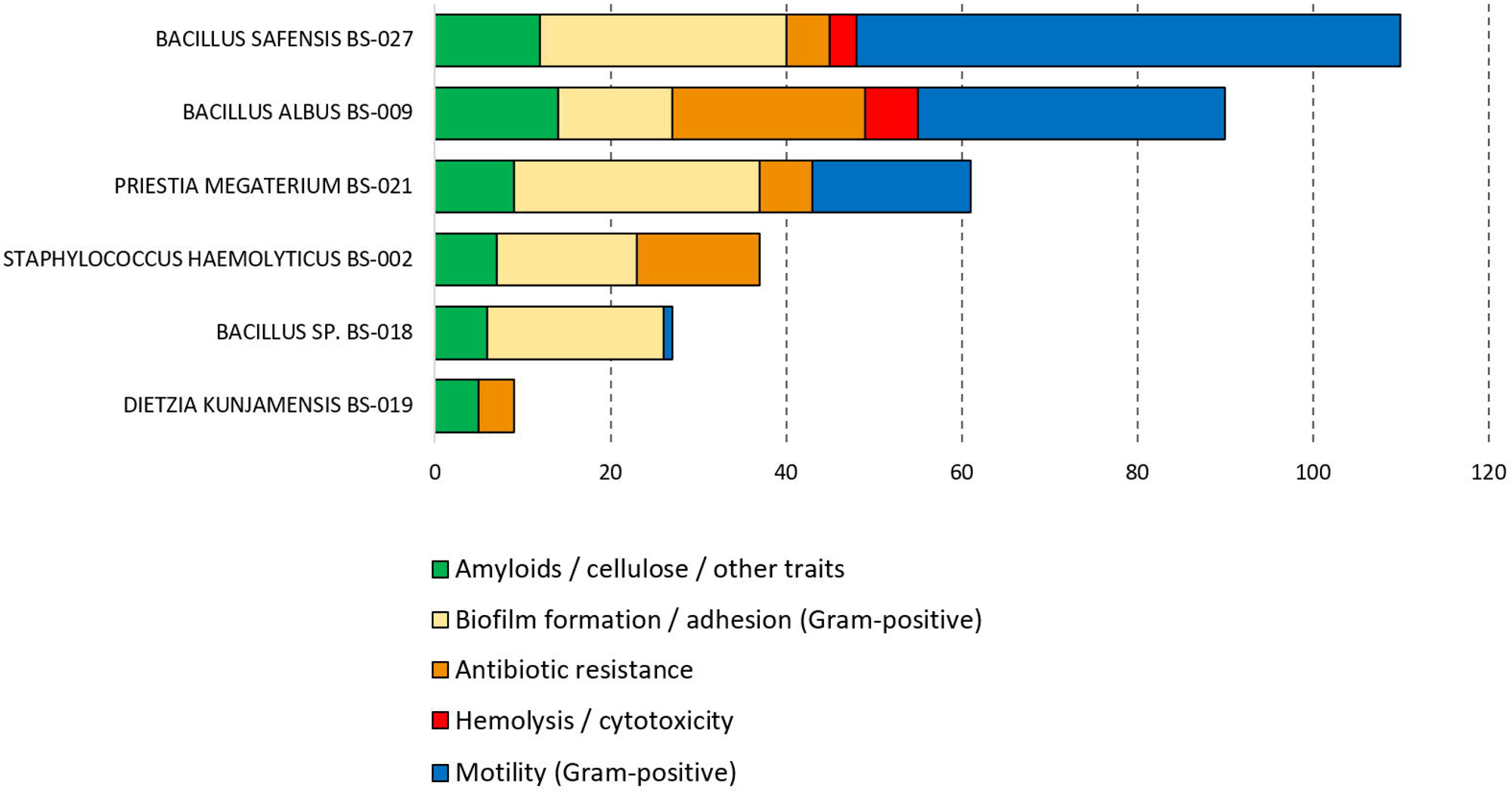
Broad functional annotation profiles of six representative blood-bank surface isolates. Stacked bars show the number of annotated gene hits grouped into major persistence- and virulence-associated functional categories for *Bacillus safensis* BS-027, *Bacillus albus* BS-009, *Priestia megaterium* BS-021, *Staphylococcus haemolyticus* BS-002, *Bacillus* sp. BS-018, and *Dietzia kunjamensis* BS-019. Functional categories include amyloid/cellulose and related traits, biofilm formation and adhesion in Gram-positive bacteria, antibiotic resistance, hemolysis/cytotoxicity, and motility-associated functions. The profiles highlight the larger functional repertoires of *B. safensis* BS-027 and *B. albus* BS-009, particularly for motility and resistance-associated annotations, and the comparatively reduced number of recovered target annotations in *D. kunjamensis* BS-019.

**Figure 7.**
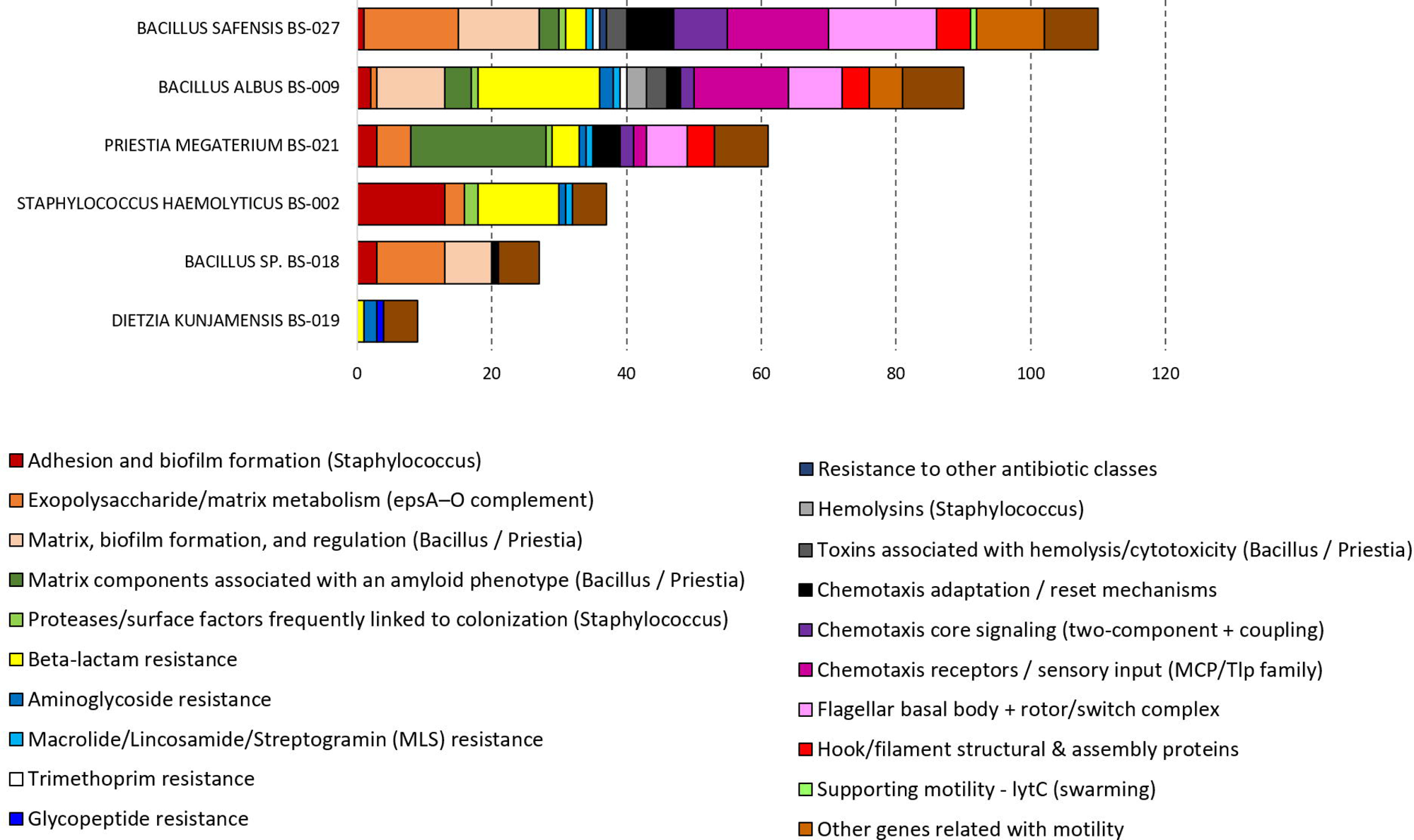
Subcategory-level distribution of persistence-, resistance-, cytotoxicity-, and motility-associated annotations in six sequenced isolates. Stacked bars show the number of annotated gene hits assigned to functional subcategories related to adhesion and biofilm formation, exopolysaccharide and matrix metabolism, amyloid-associated matrix components, antibiotic resistance classes, hemolysins and cytotoxicity-associated toxins, chemotaxis, flagellar structure and assembly, and other motility-related genes. The distribution reveals strain-specific functional patterns, including enrichment of chemotaxis, flagellar, and matrix-associated annotations in *B. safensis* BS-027, a broad resistance and toxin-associated repertoire in *B. albus* BS-009, and adhesion/biofilm-associated determinants in *S. haemolyticus* BS-002.

**Figure 8.**
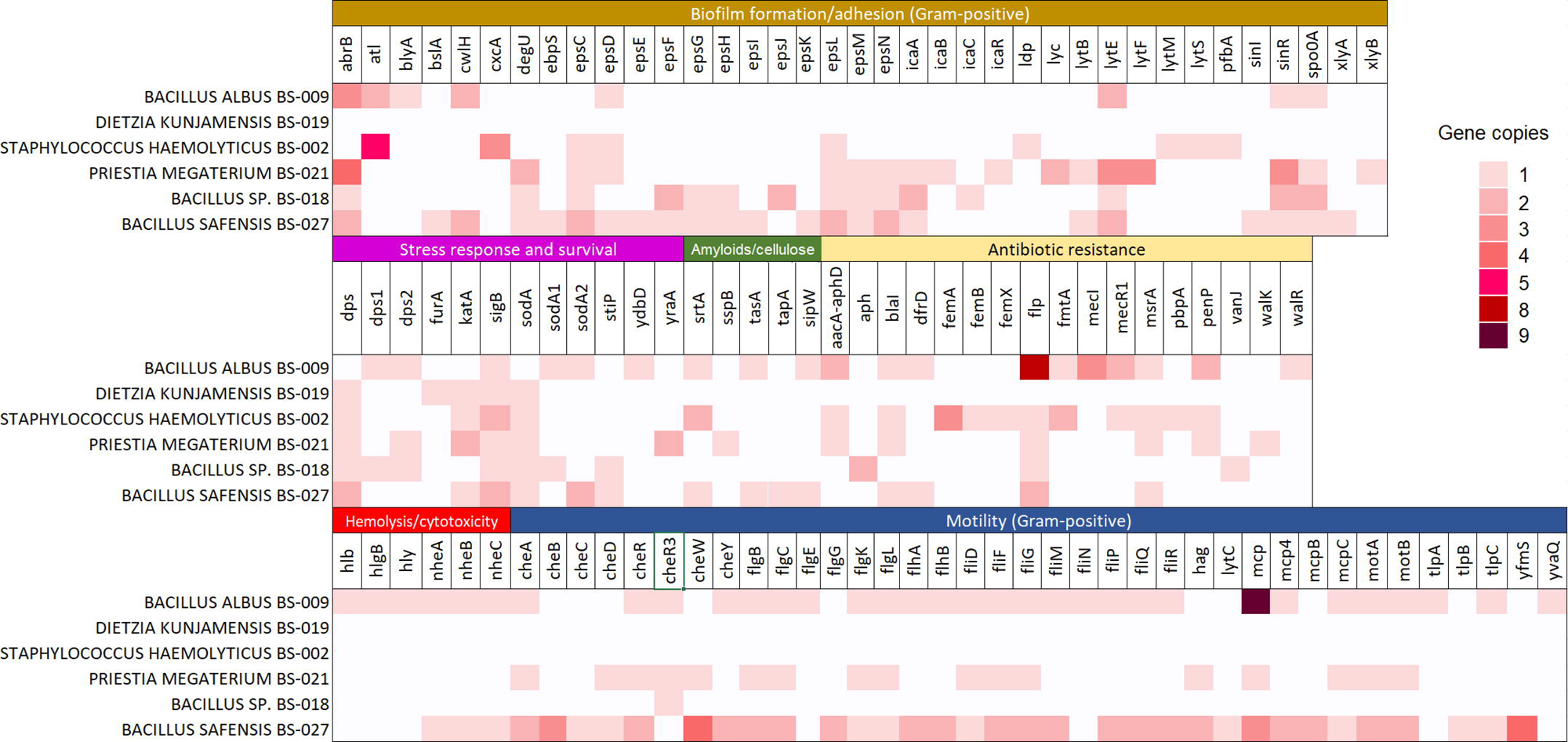
Gene-level heatmap of selected functional determinants associated with surface persistence and potential virulence in six representative isolates. Rows correspond to sequenced strains and columns correspond to target genes grouped by functional module: biofilm formation/adhesion in Gram-positive bacteria, stress response and survival, amyloid/cellulose production, antibiotic resistance, hemolysis/cytotoxicity, and motility-associated functions. Color intensity indicates the number of gene copies recovered for each target. The heatmap shows strain-specific signatures, including abundant motility-related genes in *B. safensis* BS-027, resistance-related and hemolysis/cytotoxicity-associated determinants in *B. albus* BS-009, adhesion and cell-envelope-associated loci in *S. haemolyticus* BS-002, and a limited number of recovered target annotations in *D. kunjamensis* BS-019 despite its strong biofilm-forming and motility phenotypes.

More detailed analysis at the functional subcategory level revealed pronounced differences in the distribution of genes associated with adhesion, extracellular matrix synthesis, chemotaxis, flagellar assembly, antibiotic resistance, and hemolytic activity (Fig. 7). Particularly notable was the enrichment of chemotaxis- and flagellum-related genes in *B. safensis* BS-027, whereas *B. albus* BS-009 displayed a broader repertoire of resistance- and toxin-associated genes. Conversely, *S. haemolyticus* BS-002 showed a predominance of genes involved in adhesion and Gram-positive biofilm formation, consistent with its ecological adaptation to persistent surface colonization. The integration of these functional categories with the corresponding phenotypic and ultrastructural observations is presented in Table 5, highlighting the distinct persistence strategies adopted by each representative isolate.

Comparison of genomic predictions with phenotypic characterization demonstrated that several persistence-associated phenotypes were supported by corresponding functional gene repertoires (Fig. 8). Isolates exhibiting high motility generally possessed expanded sets of genes involved in chemotaxis and flagellar assembly, whereas strong biofilm producers frequently contained genes associated with adhesion, extracellular matrix production, and stress tolerance. Likewise, multidrug-resistant isolates harbored diverse antimicrobial resistance determinants consistent with their susceptibility profiles.

Nevertheless, important genotype–phenotype discrepancies were also observed. *Dietzia kunjamensis* BS-019 displayed one of the strongest biofilm-forming phenotypes despite harboring a relatively limited number of annotated persistence-related genes. Similarly, BS-018 exhibited α-hemolysis despite lacking obvious homologues of well-characterized toxin-associated determinants, suggesting that additional regulatory pathways or poorly annotated genes may contribute to this phenotype. These observations indicate that bacterial persistence in blood bank environments cannot be explained solely by gene content but likely reflects complex interactions among regulatory networks, gene expression, and environmental adaptation.

The integrated analysis summarized in Table 5 illustrates that no single persistence profile was shared among all isolates. Instead, each strain combined distinct structural, phenotypic, and genomic features, defining different ecological persistence strategies. For example, BS-027 was characterized as a highly adapted surface colonizer with extensive motility- and biofilm-associated determinants, whereas BS-019 behaved as a biofilm specialist despite possessing a comparatively reduced repertoire of annotated persistence genes. Similarly, BS-002 primarily relied on adhesion and surface colonization mechanisms, while BS-009 combined multidrug resistance, motility, and toxin-associated genes, suggesting greater opportunistic potential. BS-021 exhibited a stress-tolerant environmental survival strategy dominated by sporulation- and stress-response genes, whereas BS-018 may represent a novel lineage whose persistence mechanisms remain largely unexplored.

Collectively, the integration of ultrastructural imaging, phenotypic characterization, and comparative genomics demonstrates that bacterial persistence on blood bank surfaces emerges from multiple complementary adaptive strategies rather than from a single conserved genetic signature. This integrative framework reveals distinct ecological persistence profiles among the recovered isolates and highlights the value of multimodal approaches for environmental surveillance in healthcare-associated settings.

## DISCUSSION

The present study demonstrates that blood bank surfaces should not be regarded merely as passive sites of environmental contamination but rather as selective ecological niches that favor the establishment of viable microorganisms possessing complementary persistence-associated traits. The culturable microbiome recovered from these environments was dominated by environmentally derived Gram-positive bacteria, particularly members of the genera *Bacillus*, *Micrococcus*, *Dietzia*, *Kocuria*, *Brevibacterium*, and *Lysinibacillus*. These microorganisms are widely distributed in air, dust, ventilation systems, and indoor built environments and are recognized for their ability to tolerate desiccation, oxidative stress, nutrient limitation, and recurrent sanitation procedures (Qiu et al., 2022; Bitew et al., 2021; Vestby et al., 2020; Ramirez-Arcos et al., 2023; Shiota and Nakayama, 1997; Kooken et al., 2012; Madsen et al., 2023).

However, these compositional patterns should be interpreted within the methodological framework of the present study. Because the analyses were intentionally based on culturable microorganisms recovered on LB medium at 30 °C, the observed community represents only the viable fraction capable of growing under these experimental conditions. Fastidious, slow-growing, stressed, or viable-but-nonculturable microorganisms—including numerous Gram-negative taxa—were likely underrepresented. Rather than describing the complete blood bank microbiome, this approach specifically targeted microorganisms capable of surviving environmental conditions and allowed their subsequent phenotypic and genomic characterization. This strategy is particularly relevant because viable surface-associated bacteria are those most likely to participate in biofilm formation, environmental persistence, and potential contamination events (Vestby et al., 2020).

The predominance of airborne environmental taxa further supports the hypothesis that ecological filtering, rather than direct contamination from blood products, shapes microbial communities inhabiting blood bank surfaces. For example, *Micrococcus luteus*, the predominant species recovered from the molecular biology laboratory is frequently associated with airborne dissemination, human skin microbiota, and indoor environments, where it exhibits remarkable resistance to ultraviolet radiation and desiccation (Shiota and Nakayama, 1997; Kooken et al., 2012; Madsen et al., 2023). Likewise, members of the *Bacillus* sensu lato group—including *B. altitudinis*/pumilus, *B. albus*, *B. safensis*, and *Priestia megaterium*—are ubiquitous spore-forming bacteria commonly detected in aerosols, dust particles, and ventilation systems, enabling long-term persistence under fluctuating environmental conditions (Qiu et al., 2022; Madsen et al., 2023).

These observations are consistent with recent international surveys demonstrating that environmental microbiological monitoring remains inconsistently implemented throughout blood transfusion facilities despite increasing awareness of its importance for transfusion safety (Ramirez-Arcos et al., 2023). Collectively, our findings support the concept that blood bank surfaces represent dynamic microbial ecosystems in which environmental selection favors microorganisms capable of surviving repeated cleaning cycles and prolonged exposure to abiotic stress.

One of the principal findings of this study is that bacterial persistence on blood bank surfaces appears to result from the coexistence of multiple adaptive mechanisms rather than from a single virulence-associated trait. Phenotypic analyses demonstrated that numerous isolates simultaneously expressed biofilm formation, extracellular matrix production, motility, hemolytic activity, and multidrug resistance, indicating that persistence represents a multifactorial ecological strategy shared by phylogenetically diverse microorganisms. Indeed, biofilm formation emerged as one of the most relevant persistence-associated phenotypes identified in the present study. Numerous isolates belonging to *Bacillus*, *Burkholderia*, *Dietzia*, and *Staphylococcus* developed mature biofilms, while scanning electron microscopy revealed structured microbial communities embedded within abundant extracellular polymeric substances on representative blood bank surfaces. These observations provide direct structural evidence that colonization occurs predominantly as organized biofilms rather than isolated planktonic cells. Such biofilm architecture is known to enhance bacterial survival by increasing tolerance to disinfectants, desiccation, nutrient limitation, and antimicrobial exposure (Mishra et al., 2024; Taner et al., 2024; Abdallah et al., 2014; Li et al., 2024). As proposed by Flemming and Wingender (2010), the extracellular matrix functions as a protective and interactive scaffold that promotes both bacterial survival and microbial cooperation.

Similarly, Congo Red-positive colony morphotypes consistent with extracellular matrix production were widespread among the recovered isolates. Although this assay provides only indirect evidence of amyloid-like fibers and cellulose biosynthesis, the observed colony morphologies strongly suggest the capacity to produce surface-associated extracellular structures involved in biofilm maturation (Romling et al., 2000; Coleri et al., 2017; Romero et al., 2010). The diversity of morphotypes observed among phylogenetically unrelated bacteria indicates that matrix production represents a convergent adaptive strategy rather than a taxon-specific characteristic. Motility represented an additional persistence-associated phenotype. Highly motile isolates, particularly *Bacillus* species, are likely to possess an enhanced capacity for surface exploration, colonization, and subsequent biofilm establishment. Together with biofilm formation and extracellular matrix production, motility probably facilitates continuous recolonization of environmental surfaces following routine sanitation procedures.

Antimicrobial resistance further reinforced this persistence model. More than 30% of the recovered isolates fulfilled the criteria for multidrug resistance, including both Gram-positive and Gram-negative bacteria. Particularly noteworthy were *Staphylococcus haemolyticus*, *Burkholderia cenocepacia*, and *Stutzerimonas stutzeri*, which combined MDR phenotypes with biofilm formation and motility. These findings agree with increasing evidence indicating that healthcare environments function as environmental reservoirs of antimicrobial resistance beyond patient-associated niches (Zhuang et al., 2021; Frieri et al., 2017; Bowler et al., 2020; Sharma et al., 2026; Venkatesan et al., 2015; Eltwisy et al., 2022; Mohammedali and Abed, 2025; LeChevallier et al., 2024; Fazli et al., 2014; Alcàcer-Almansa et al., 2025). Rather than acting independently, antimicrobial resistance, biofilm formation, extracellular matrix production, and motility appear to operate as complementary mechanisms enhancing long-term bacterial persistence on abiotic surfaces.

Whole-genome sequencing substantially expanded the ecological interpretation of the phenotypic observations by demonstrating that persistence-associated traits are largely strain-dependent rather than uniformly distributed across bacterial taxa. Instead of revealing a single conserved persistence genotype, comparative genomics identified distinct combinations of functional determinants associated with adhesion, extracellular matrix production, chemotaxis, stress tolerance, antimicrobial resistance, and cytotoxicity. *Bacillus albus* BS-009 and *Bacillus safensis* BS-027 exhibited the broadest genomic repertoires associated with persistence. BS-009 combined numerous resistance determinants with toxin-associated genes and an expanded chemotaxis and flagellar module, whereas BS-027 was enriched in genes related to motility, extracellular matrix production, and biofilm formation. Conversely, *Staphylococcus haemolyticus* BS-002 concentrated adhesion-associated loci together with antimicrobial resistance determinants, supporting its capacity to persist on healthcare-associated surfaces.

One particularly relevant finding was the identification of BS-018, whose average nucleotide identity remained below the accepted species threshold of 95% (Richter and Rosselló-Móra, 2009). This observation suggests the presence of a potentially undescribed environmental lineage colonizing blood bank surfaces and emphasizes how little is currently known about microbial diversity in built healthcare environments.

An equally important observation was the frequent discordance between genotype and phenotype. Several strains carrying numerous biofilm-associated genes failed to produce biofilms under the tested conditions, whereas other isolates displayed strong persistence-associated phenotypes despite possessing relatively few annotated determinants. The remarkable biofilm-forming ability of *Dietzia kunjamensis* BS-019 despite its comparatively limited genomic repertoire illustrates this phenomenon. Such discrepancies likely reflect regulatory complexity, environmental modulation of gene expression, and the still incomplete annotation of environmental bacterial genomes (Orkara et al., 2026; Deka et al., 2026; Mokari et al., 2026).

Rather than representing inconsistencies, these genotype–phenotype differences reinforce the importance of integrating comparative genomics with phenotypic assays and bioimaging. Together, these complementary approaches demonstrate that bacterial persistence cannot be reliably inferred from genomic information alone but instead emerges from complex interactions among genetic potential, physiological regulation, and environmental conditions.

The ecological perspective emerging from this study has important implications for transfusion medicine. Rather than viewing environmental contamination exclusively as an occasional introduction of microorganisms from blood products or personnel, our findings suggest that blood bank facilities harbor resident microbial communities capable of long-term persistence on critical processing surfaces. This interpretation agrees closely with recent recommendations from the ISBT Transfusion-Transmitted Infectious Diseases Working Party, which emphasized the urgent need for standardized environmental microbiological monitoring throughout the blood supply chain (Ramirez-Arcos et al., 2023; Ramirez-Arcos et al., 2015; Blajchman, 2004; Brecher and Hay, 2005; Cloutier et al., 2022; García-Otálora et al., 2024). The repeated recovery of structured biofilm-forming communities from platelet agitators, centrifuges, worktables, and air-conditioning systems suggests that routine sanitation alone may be insufficient to eliminate persistent microorganisms completely. Importantly, the integration of scanning electron microscopy, phenotypic characterization, and whole-genome sequencing provided a multidimensional framework for identifying microorganisms with greater persistence potential. Beyond simple taxonomic identification, this multimodal approach allowed structural organization, adaptive phenotypes, and genomic determinants to be interpreted together, providing a more realistic assessment of environmental biosafety risks. These findings support the incorporation of integrated environmental surveillance programs combining microbiological monitoring, bioimaging, and genome-resolved analyses to optimize infection prevention strategies within transfusion services.

This study has several limitations. Sampling was restricted to a single time point and relied primarily on culture-dependent isolation using LB medium at 30 °C, conditions that selectively recover only the viable and culturable fraction of the microbial community. Consequently, fastidious microorganisms, viable-but-nonculturable bacteria, and organisms requiring alternative growth conditions were probably underestimated. Furthermore, genomic analyses were limited to six representative isolates, and no culture-independent community profiling or transcriptomic analyses were performed.

Future investigations should combine longitudinal environmental sampling with quantitative microbiological approaches, 16S rRNA gene sequencing, shotgun metagenomics, transcriptomics, and spatially resolved imaging techniques to characterize both the taxonomic composition and functional activity of blood bank surface microbiomes. Such integrative approaches will improve our understanding of the ecological processes governing microbial persistence and support the development of evidence-based environmental monitoring strategies for transfusion medicine.

## Data Availability

All data produced in the present study are available upon reasonable request to the authors

## Credit authorship contribution statement

V.A. had the original project idea and designed the research study; C.D., G.M. and V.A. performed the sampling; S.G. and N.A. performed the isolation; C.D. and M.G. performed the phenotypic characterization; C.D. and L.M. processed samples and obtained the bioimages. D.A. analysed the genomic data; V.A., C.D. and D.A. wrote the manuscript; V.A. and M.G. contributed essential reagents and equipment; V.A. supervised the research, obtained funding and reviewed and edited the final version of the manuscript. All authors read and reviewed the final version of the manuscript.

## Funding

This work was funded by the following projects of Universidad San Pablo-Tucumán: ISYCAV-1222-2025; ISYCAV-1119-2024; ISYCAV-1212-2023; ISYCAV-841-2019; ISYCAV-842-2019; and PIUNT G-603-2017 from SCAIT - Universidad Nacional de Tucumán. V.A. and M.G. are staff researchers and L.M. and S.G. are staff technicians of the National Council of Research and Technology (CONICET). C. D. and N.A. were recipients of national research scholarships from CONICET. VHA was a recipient of a Georg Foster Scholarship for Experienced Researchers, Alexander von Humboldt Foundation (2021-2023) and of a grant of the Williams Foundation (Fondos Complementarios para Proyectos con Impacto en el Territorio 2024).

## Declaration of competing interest

The authors have declared that no competing interests exist.

## Acknowledgments

We thank Dr. Felicitas Agote, Director of the Banco Central de Sangre de Tucumán “Dr. César Guerra” (SiPROSA), for kindly granting authorization and access to sampling. We also would like to acknowledge Biochemists Julieta Amado and José Sánchez for their contributions in the interpretation of the antibiograms, as well as the Mussa Infectious Diseases Center, where this part of the work was carried out. We would also like to thank technical assistance of Hernán Esquivel and Roberto Fanjul for imaging samples. All micrographs were taken at the Centro Integral de Microscopía Electrónica (CIME), belonging to UNT and CONICET, in Tucumán, Argentina.

## Notes

### Competing Interest Statement

The authors have declared no competing interest.

